# Discrepancies Between Self-Reported and High-Precision UWB-Measured Close Contacts: Evidence from a Japanese Junior High School

**DOI:** 10.1101/2025.09.11.25335361

**Authors:** Hayato Shinto, Kenji Mizumoto, Yu Ohki, Gerado Chowell, Kei Saito, Yoshihiro Takayama

**Author notes:** **Address for correspondence:** Kenji Mizumoto, Graduate School of Advanced Integrated Studies in Human Survivability, Kyoto University, Yoshida-Nakaadachi-cho, Sakyo-ku, Kyoto 606-8306, Japan. These authors contributed equally to this work (Hayato Shinto, Kenji Mizumoto).

## Abstract

**Background:** Accurate identification of close contacts is essential for effective infection control. However, school-based contact tracing typically relies on retrospective self-reports, which are prone to recall bias. Ultra-wideband (UWB) indoor positioning offers centimeter-level spatial resolution but has rarely been applied in educational settings. This study is the first to benchmark self-reported “close contacts” against centimeter-level ultra-wideband (UWB) positioning in a real classroom over multiple consecutive days, extending beyond prior RFID- or Bluetooth-based school studies.

**Methods:** We conducted a five-day observational study in a junior high school classroom in Japan (36 participants; 33 with complete questionnaire and UWB data). Close contacts were identified via self-reported questionnaires and UWB positioning under multiple distance–time thresholds, with 1.0 meter for ≥15 minutes as the official baseline. Discrepancies were quantified using participant-level consistency rates, and sensitivity analyses evaluated alternative thresholds.

**Findings:** Discrepancies between the two methods were consistently large under the 1.0 m/15 min definition. The median discrepancy was –0.16, indicating systematic overreporting in questionnaires. The smallest discrepancies occurred at 1.5 m for ≥10 min, where UWB and recall most closely aligned. Notably, UWB continued to identify new unique contacts throughout the week, whereas questionnaire-based reporting plateaued after three days. Higher confidence in self-reports was paradoxically associated with greater overreporting. Teacher–student reciprocity was absent, despite the teacher reporting contact with all students.

**Interpretation:** In a fixed-seating classroom, self-reported and UWB-measured close contacts diverged substantially. By leveraging centimeter-precision UWB tracking and threshold calibration, our study provides the first empirical evidence that current operational definitions reflect memory alignment more than actual spatial interactions. Incorporating UWB positioning and empirically optimized criteria may improve the accuracy and feasibility of contact tracing in schools, while minimizing unnecessary quarantines and educational disruption.

**Funding:** Japan Society for the Promotion of Science

**Research in context:** *Evidence before this study:* We searched PubMed and Google Scholar from Jan 1, 2000, to July 31, 2025, using the terms “close contact”, “self-report”, “recall bias”, “school”, “Bluetooth”, “RFID”, and “ultra-wideband (UWB)”. Previous school-based studies relied on diaries, Bluetooth, or RFID sensors, which provide only meter-level accuracy and showed large discrepancies between reported and measured contacts. To our knowledge, no study had systematically applied UWB, with centimeter-level precision, to evaluate close contacts in a real-world classroom setting.

*Added value of this study:* To our knowledge, this is the first study to compare self-reported close contacts with UWB-measured proximity in a junior high school classroom over five consecutive days. We show that self-reports consistently overestimated the number of close contacts compared with UWB, that recall plateaued after three days, and that the smallest discrepancies were observed under a threshold of 1.5 m for ≥10 min rather than the official 1.0 m for ≥15 min definition. These findings demonstrate that operational definitions align more with recall patterns than with actual spatial interactions.

*Implications of all the available evidence:* Our findings suggest that current close contact definitions may lead to substantial misclassification in school settings, resulting in unnecessary quarantines and educational disruption. Incorporating UWB or similar high-precision positioning technologies, together with empirically optimized thresholds, could improve the accuracy and feasibility of school-based contact tracing and help balance infection control with educational continuity.

## Introduction

During outbreaks of emerging infectious diseases, the operational definition of a “close contact” determines who is quarantined, tested, or monitored. Inaccurate or inconsistently applied definitions risk both missing high-risk exposures and imposing unnecessary restrictions. For COVID-19, the U.S. Centers for Disease Control and Prevention (CDC) defined a close contact as being within approximately 1.8 meter of an infected person for a cumulative ≥15 min during the infectious period [1], consistent with evidence from a systematic review showing that physical distancing of at least 1–2 m substantially reduces transmission risk [2]. In Japan, the definition evolved from “within 2 m” with no time criterion to “within 1 m for ≥15 min” regardless of mask use, with quarantine durations shortened over time [3–5].

In practice, close contacts are typically identified via retrospective self-reports, which are prone to recall bias, overreporting, and underreporting [6,7]. Earlier studies in healthcare and long-term care settings have compared self-reports with Bluetooth beacon or RFID sensor data, revealing substantial discrepancies [8,9]. While Bluetooth and RFID systems offer spatial resolutions of 1–3 m, their limited accuracy constrains their utility in evaluating the validity of close contact definitions [10,11]. School-based sensor studies in the U.S., France, and Italy have also demonstrated systematic differences between diaries and RFID-based proximity data [12–14], but these approaches lacked the centimeter-level precision required to rigorously test operational thresholds. In contrast, ultra-wideband (UWB) positioning achieves accuracies on the order of 10–30 centimeters [15–17], enabling high-resolution tracking of interpersonal distance and duration and offering a new opportunity to reassess close contact definitions in schools.

Despite its technical potential, UWB has seen limited use in infectious disease control research. Although widely adopted in logistics, sports, and industrial environments, its application in schools and healthcare remains rare—largely due to infrastructure requirements and cost. Recent studies have demonstrated the feasibility of using UWB to monitor or visualize interpersonal interactions in organizational settings [16,18], but to date, no study has systematically applied UWB to quantify discrepancies between self-reports and high-precision measurements in classrooms.

A retrospective study in elderly care facilities in Okinawa Prefecture, Japan, found that outbreak sizes were significantly smaller when testing was initiated based on reported contact history, compared to routine weekly screening of staff [19]. This finding underscores the importance of accurate and timely contact identification in outbreak control. Yet, the degree to which self-reported contacts in schools align with actual spatial interactions has remained untested with centimeter-level methods.

Here, we present the first classroom-based study to directly compare self-reported close contacts with UWB-measured proximity data over five consecutive school days. By combining questionnaire-based recall with high-resolution positioning, we assess the extent and patterns of discrepancies under multiple distance–time thresholds, including the official 1.0 m/15 min definition. We further identify an empirically optimized threshold (1.5 m/10 min) that minimized diary–device discrepancies, providing new evidence for how operational definitions align with recall and measurement in practice.

## Methods

This study was designed to directly compare close contacts identified through self-reported questionnaires with those detected using high-precision UWB positioning, and to quantify discrepancies between the two methods.

### Study design and setting

This observational study was conducted in January 2025 at a junior high school in Wakayama Prefecture, Japan. The school has one class per grade; the survey focused on the movements of students within a single second-grade classroom. The classroom measures 7.7 m in depth and 7.4 m in width (total floor area 57.0 m²) with a ceiling height of 3 m, dimensions representative of standard Japanese classrooms.

In Japan, junior high school covers three grades, typically attended by students aged 12 to 15 years [20]. Schools operate on a five-day school week (Monday to Friday), with Saturday classes largely phased out in public schools [21]. Students are assigned to fixed homerooms for most classes, and school lunches are eaten in the classroom. At the study site, the school day runs from 08:25 to 15:40 and includes six 50-minute lessons, with lunch from 12:00 to 13:00.

The survey was conducted over five consecutive weekdays (Monday through Friday), covering the entire homeroom period each day (08:25–15:40). The study area was limited to the classroom. UWB tags were distributed and collected every day by the homeroom teacher at the start and end of the survey. Eight receivers were installed across the classroom.

Lessons such as music, physical education, art, and science experiments were held outside the homeroom and, therefore, excluded. The number of homeroom-based lessons during the survey period was five on Monday (Day 1), five on Tuesday (Day 2), one on Wednesday (Day 3), four on Thursday (Day 4), and five on Friday (Day 5).

### Participants

Thirty-eight individuals were eligible for the study (37 students and one teacher). Of these, 36 participants (35 students and one teacher; 94.7%) provided written informed consent and carried UWB tags during the survey. Questionnaire responses were obtained from 33 participants, and analyses were restricted to these respondents.

### Ethics approval and consent to participate

The study protocol was reviewed and approved by the Ethics Committee of Kyoto University Graduate School of Medicine (Approval No. R6-G-8-2). Written informed consent was obtained from all participants and their legal guardians prior to participation.

### Questionnaire survey

At the end of the survey period, each participant completed a structured questionnaire (Additional file 2) developed for this study. Items included attendance number, grade, sex, age, role (student or teacher), daily list of close contacts, and confidence level in recall (5-point Likert scale). For the purpose of the questionnaire, a close contact was defined as anyone who had contact with the participant within 1 meter for a cumulative total of ≥15 m during the school day, regardless of mask use.

### UWB positioning measurements

The UWB indoor positioning system comprised wearable transmitters (tags) and fixed receivers. Eight receivers were deployed within the classroom to capture three-dimensional mobility data at a frequency of 1 Hz, with a nominal measurement error of 10–30 cm [11,16]. Tags were attached to the participant’s chest to approximate torso position during interaction.

## Data processing

### Preprocessing and missing data handling

Raw data were trimmed to the study hours each day (08:23–15:40). Tag IDs were matched to participant IDs. With 36 tagged participants, 630 unique pairs were formed, and the pairwise Euclidean distance was calculated at each second. Continuous missing data ≥60 seconds were assumed to indicate the participant had left the classroom, and these periods were excluded. Periods when tags were clustered within a 50 centimeter radius in ≥10 participants (≥27% of total) were excluded to account for temporary tag collection or activities outside the classroom. These preprocessing steps were applied to the full set of 36 participants. Subsequent analyses that included questionnaire data were limited to the 33 participants who completed both components.

### Smoothing and interpolation

To improve positional accuracy, we applied Kalman filtering and a Long Short-Term Memory (LSTM) model, both of which have been validated in previous UWB positioning studies [18]. The LSTM model used a rolling input of the preceding 120 seconds to interpolate missing points and smooth coordinate fluctuations. Unlike prior work that applied smoothing to tag–anchor distances, we processed the absolute coordinate data directly.

### Contact classification

For each pair, the distance at each second was binarized: 1 if ≤1.0 m, zero otherwise. The total daily number of seconds meeting the threshold was computed. Pairs with ≥900 seconds (15 min) of contact in a day were classified as “close contacts” for that day.

### Statistical analysis

Statistical analyses were conducted using data from the 33 participants with both UWB and questionnaire data. For each distance–time threshold, close contacts were identified separately by the questionnaire and UWB methods. We quantified discrepancies between questionnaire- and UWB-identified contacts, using the consistency rate as the metric. The consistency rate was calculated for each participant on each survey day by comparing pairwise close contacts identified via questionnaire and UWB measurements. For each method, 33×33 binary matrices were created per day. Questionnaire data were symmetrized such that any reported contact in a pair was treated as mutual. The UWB matrix was subtracted from the questionnaire matrix, resulting in values of +1 (UWB only), –1 (questionnaire only), or 0 (both). The sum of these differences across each participant’s row was divided by the number of possible contacts (n = 32) to obtain the daily consistency rate. Positive values indicated underreporting in questionnaires, negative values indicated overreporting. This process was repeated for each of the five survey days, resulting in up to 165 participant-day observations. Detailed calculation procedures are described in Additional file 1 (Supplementary Methods).

## Results

As shown in Supplementary Figure S1, the histogram of one-second pairwise distances over the five-day survey exhibited a unimodal distribution with a peak around 2–3 m and a long right tail extending beyond eight meters. The overall distribution was well approximated by a Weibull distribution (see Supplementary Table S2 for parameters). Within the limited space of the classroom used in this experiment, the most common interpersonal distance was around 2–3 m, whereas a distance of about 1 meter—corresponding to the official definition of a close contact—was relatively rare.

Figure 1A and 1B compare the daily number of close contacts per participant identified by UWB positioning and self-reported in questionnaires, respectively. Full summary statistics for these daily counts are provided in Table S3 (Supplementary Materials). Over the survey period, UWB measurements detected a median of 6.0 contacts (IQR: 4.0 to 7.0) per participant, with the highest median on Day 1 (4.0, IQR: 0.0 to 5.0) and the lowest on Day 3 (0.0, IQR: 0.0 to 1.0). Questionnaire-based counts were consistently higher, with an overall median of 14.0 (IQR: 9.0 to 17.0), peaking on Day 1 (8.0, IQR: 0.5 to 11.0) and dropping to 9.0 (IQR: 5.0 to 14.8) on Day 3. Day-to-day variation was statistically significant for either method (Friedman test, p < 0.05).

**Figure 1.**
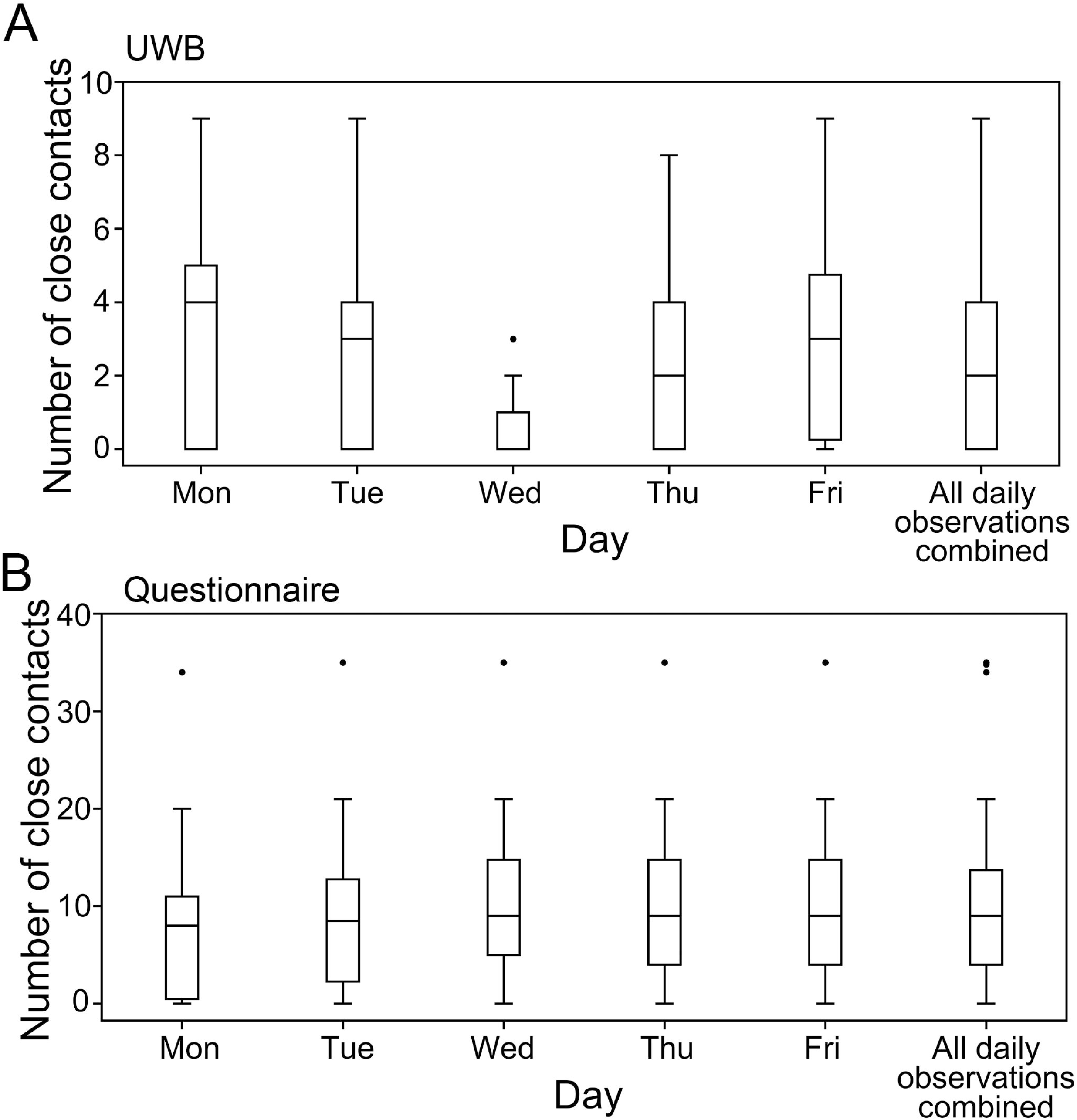
Daily number of close contacts per participant (UWB vs. Questionnaire) Close contacts were defined as individuals within 1 m for a cumulative total of ≥15 min during the school day, regardless of mask use. Each box plot represents the median, interquartile range (IQR), and statistical outliers, which are shown as black dots, for all participants (N = 33) on each survey day (Days 1–5). Days are labeled Monday through Friday, corresponding to Day 1 through Day 5 in the main text. Each boxplot shows the daily number of close contacts per participant. All daily observations combined show all close contacts (maximum 165 data points) of each participant for each day over the week. Cumulative totals over five days are not shown in this figure.

Figure 2 shows the daily discrepancies between UWB- and questionnaire-identified contacts using the baseline threshold of 1.0 m for ≥15 min [3–5]. Corresponding descriptive statistics for daily consistency rates under the official threshold are summarized in Table S4. The median discrepancy across five days was −0.16 (IQR: –0.28 to −0.05). The smallest discrepancies occurred on Day 1 (−0.11, IQR: −0.19 to 0.00) and the largest on Day 3 (−0.24, IQR: −0.36 to −0.13), with Day 3 significantly higher than Day 1 (Wilcoxon rank-sum test, p < 0.05). Positive values indicate more contacts detected by UWB (underreporting in questionnaires), whereas negative values indicate more contacts reported in questionnaires (overreporting).

**Figure 2.**
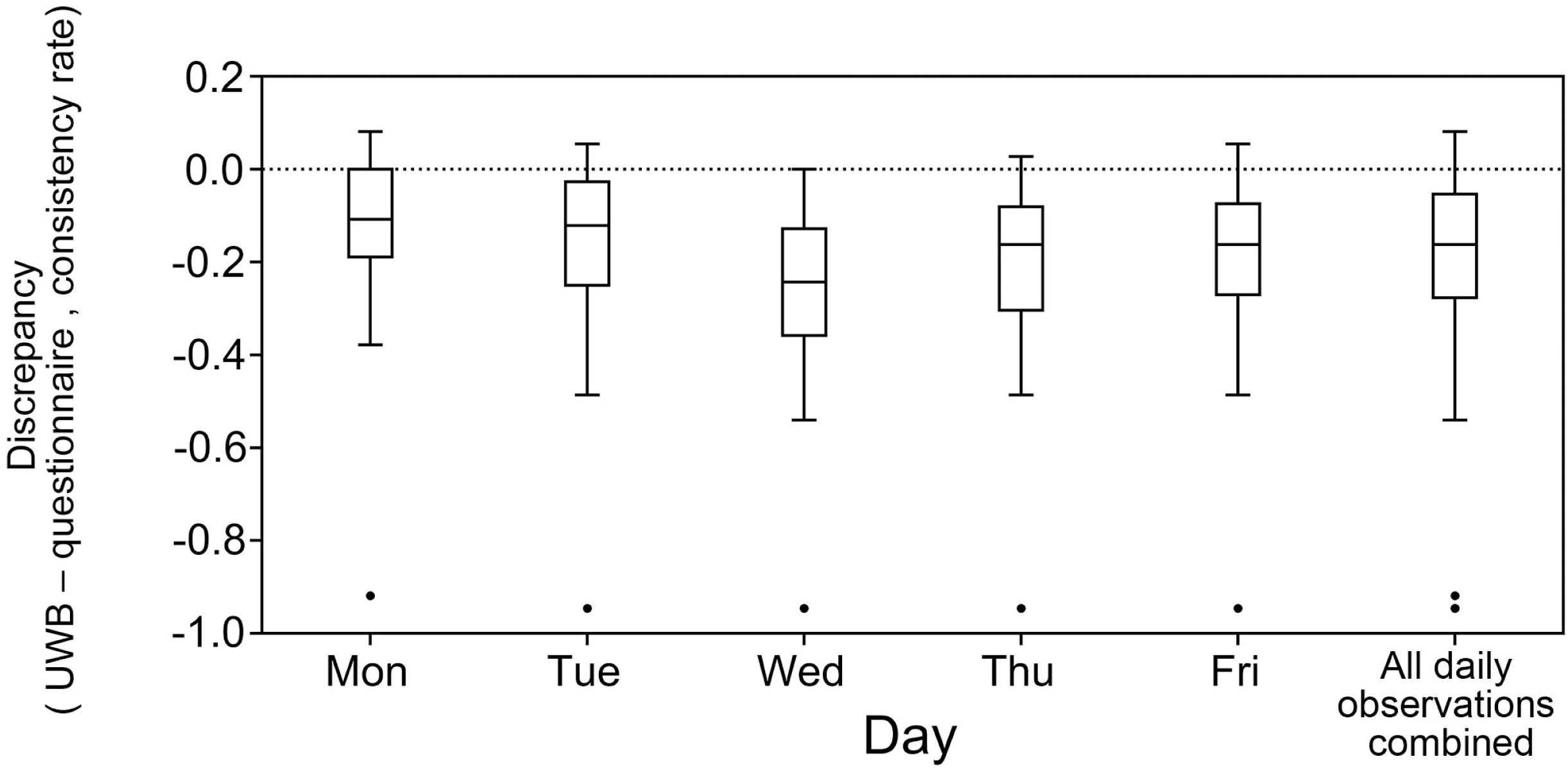
Daily discrepancies between UWB- and questionnaire-identified contacts (1.0 m, ≥15 min) Box plots show daily medians, IQRs, and statistical outliers, which are shown as black dots, across participants (N = 33). Days are labeled Monday through Friday, corresponding to Day 1 through Day 5 in the main text. Positive values indicate more contacts detected by UWB (underreporting by questionnaires), whereas negative values indicate more contacts reported in questionnaires (overreporting).

We performed sensitivity analyses to determine whether alternative thresholds could reduce discrepancies between methods. To identify the distance–time threshold that minimized discrepancies, we evaluated all 165 consistency rates (33 participants across 5 days). The smallest overall discrepancy was observed at 1.5 m for ≥10 min (Figure 3), where the median discrepancy was closest to zero, indicating balanced detection between UWB and self-reports. Among all evaluated thresholds, 1.5 m/10 min, whose median discrepancy was closest to 0, both minimized systematic bias and yielded the smallest discrepancies, suggesting that this threshold may better capture actual interaction patterns in this setting, consistent with prior evidence on recall bias [6,7] and on the utility of precise interaction mapping using UWB positioning [15,16]. Summary results across all six thresholds are provided in Table S5.

**Figure 3.**
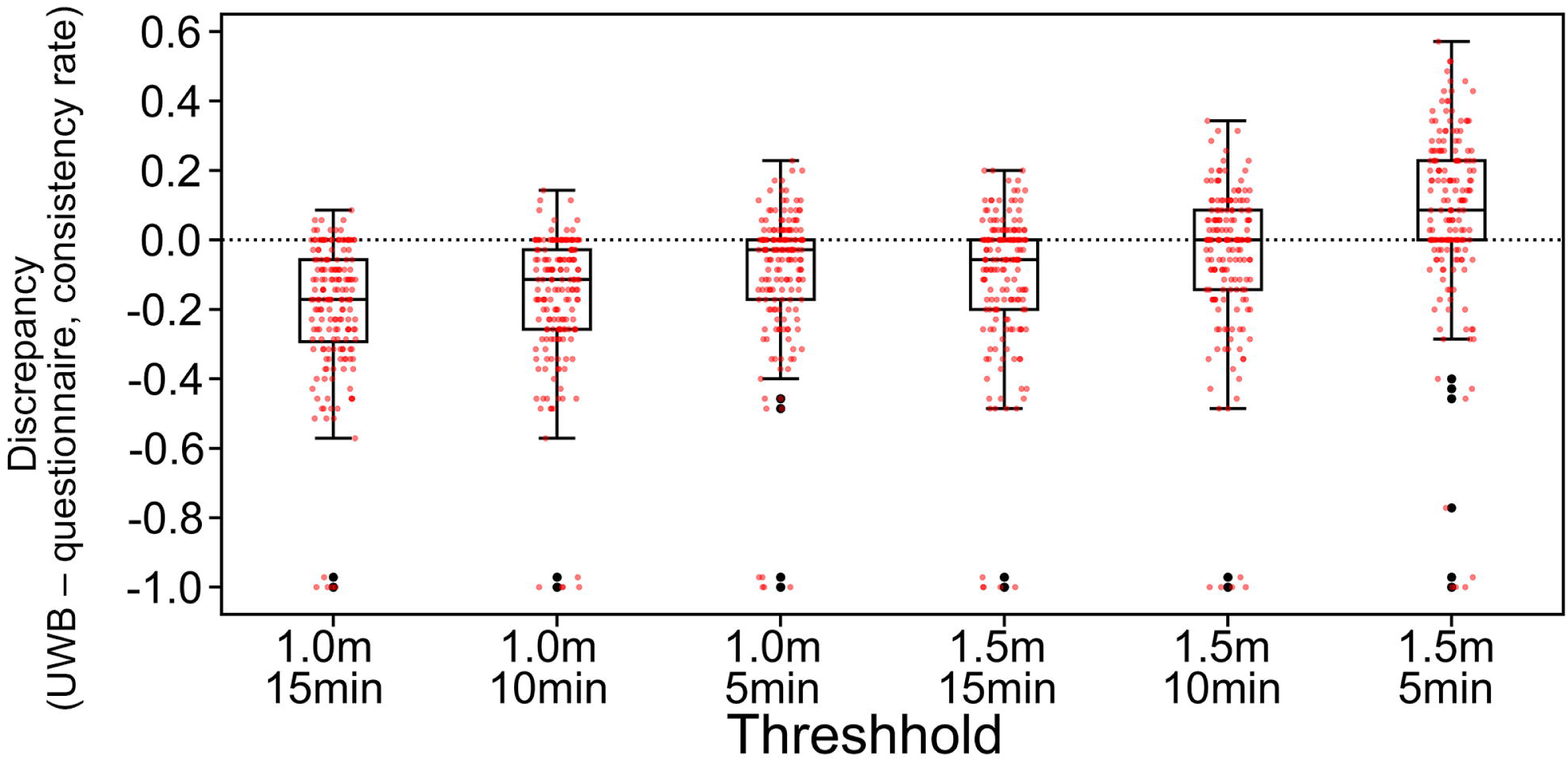
Sensitivity analysis of discrepancies under varying distance–time thresholds. Each box plot shows the distribution of discrepancies across all participant-days (33 participants × 5 days = maximum 165 observations). Positive values indicate more contacts detected by UWB, negative values indicate more contacts reported in questionnaires. Small red dots represent individual participants’ consistency rates

Applying the optimal 1.5 m/10 min threshold, Figure 4 shows median discrepancies close to zero on three of five days. Daily descriptive statistics under this optimal threshold are shown in Table S6. On Day 1, the median value was slightly positive (+0.05, IQR: 0 to +0.11), indicating a tendency toward more contacts detected by UWB than reported in questionnaires. In contrast, Day 3 showed a larger negative median (−0.16, IQR: −0.28 to −0.08), again reflecting more contacts reported in questionnaires.

**Figure 4.**
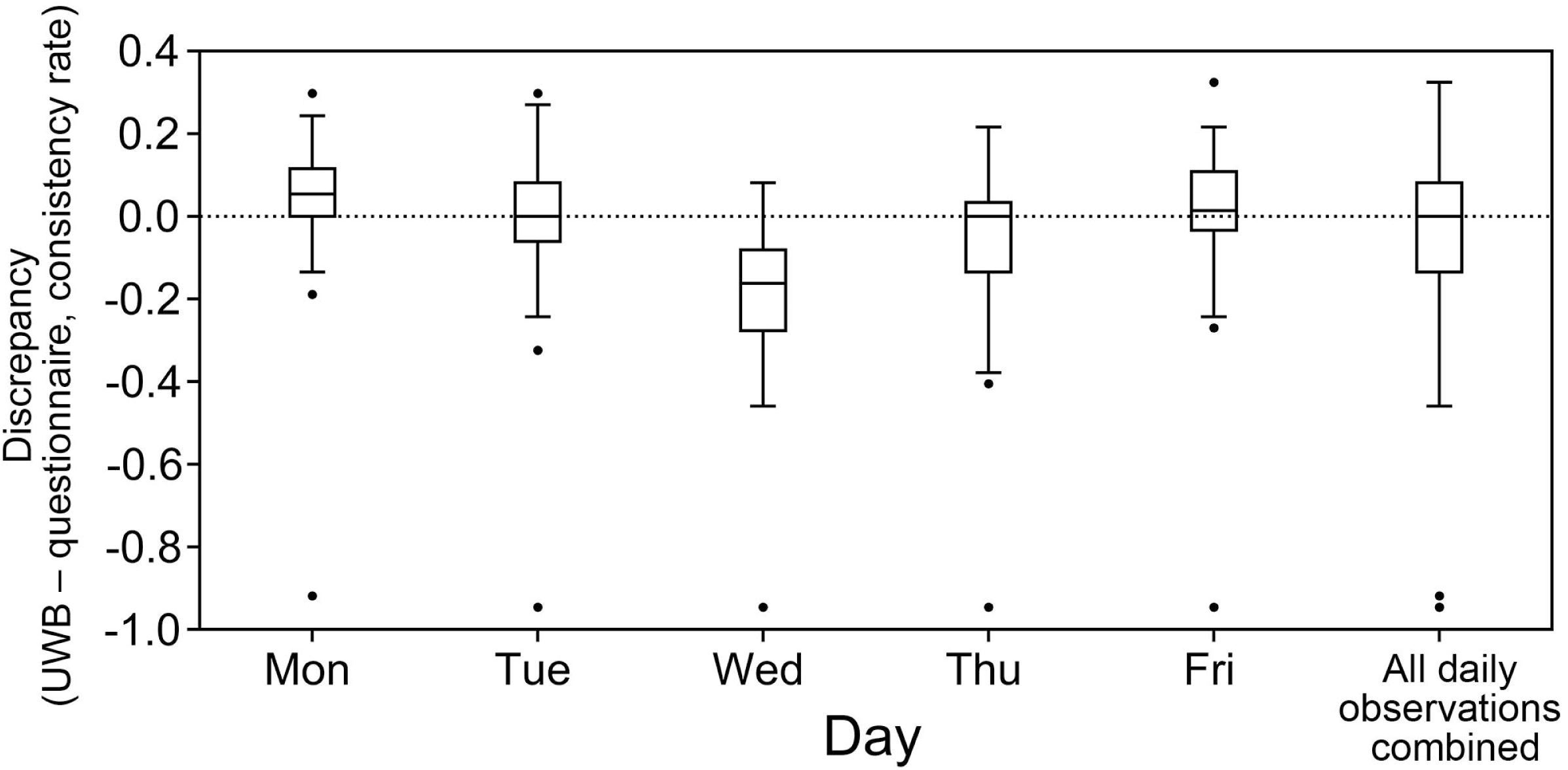
Daily discrepancies under the optimal threshold (1.5 m, ≥10 min) Daily box plots show medians, IQRs, and statistical outliers, which are shown as black dots, across participants (N = 33). Days are labeled Monday through Friday, corresponding to Day 1 through Day 5 in the main text. Values near zero indicate minimal discrepancies; positive values indicate more UWB contacts, and negative values indicate more questionnaire contacts.

Figure 5 illustrates discrepancies, stratified by participants’ confidence in their questionnaire responses. Higher confidence scores (4–5) were associated with larger negative discrepancies, indicating greater overreporting in questionnaires relative to UWB, despite participants’ perceived certainty, although variability remained high across all categories.

**Figure 5.**
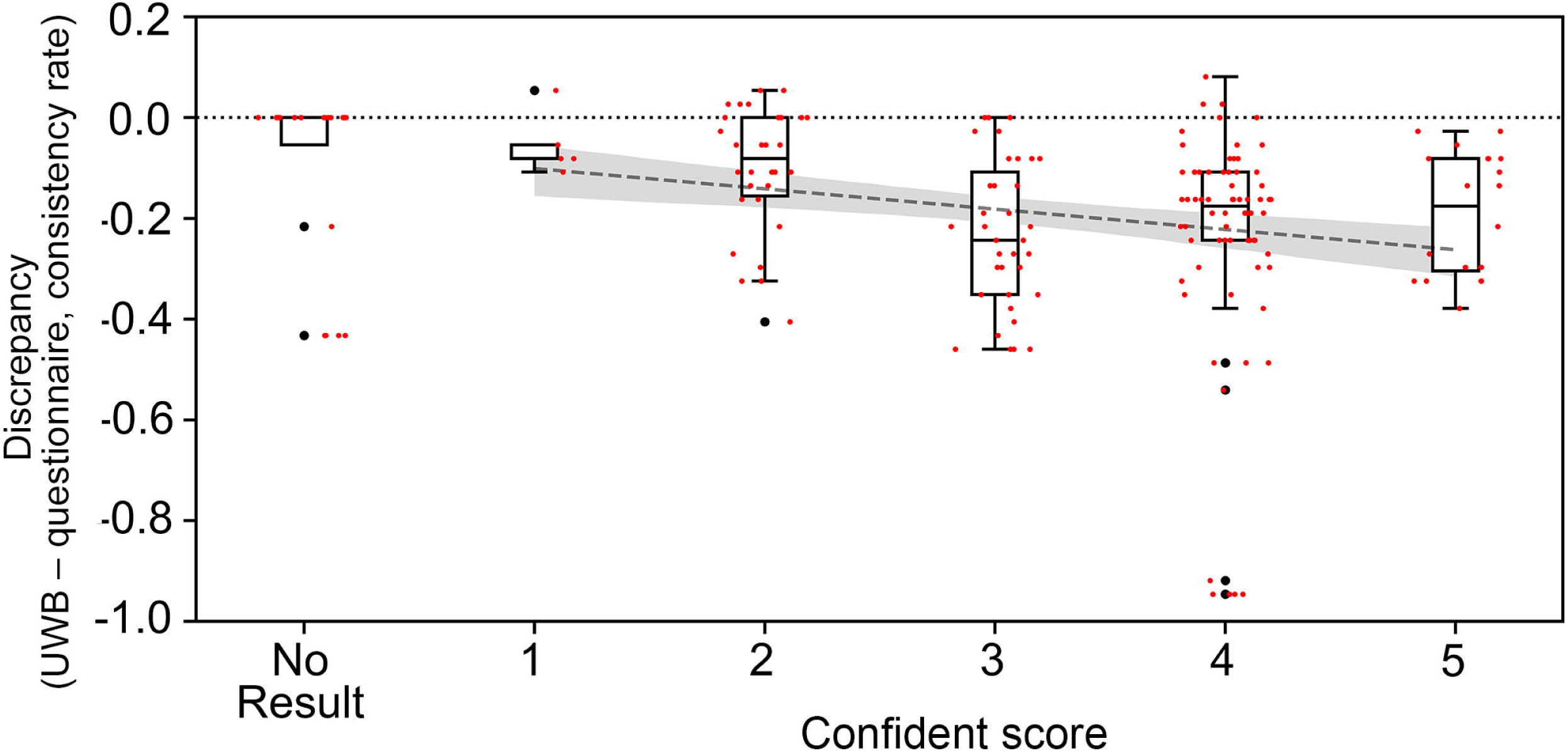
Discrepancies stratified by participants’ confidence in questionnaire responses. Confidence was rated on a 5-point Likert scale (1 = very low, 5 = very high). Box plots display medians, IQRs, and statistical outliers, which are shown as black dots, across participants (N = 33). Positive values indicate more UWB contacts, negative values indicate more questionnaire contacts. Small red dots represent individual participants’ consistency rates. The dashed line represents the fitted linear regression line. The shaded gray area indicates the 95% confidence interval around the regression line.

Additional analyses provided further context. Supplementary Figure S2 shows pairwise consistency rates based on questionnaires. While the figure itself does not indicate participant roles, our data showed that teacher–student pairs exhibited zero pairwise consistency throughout the survey period, despite the teacher reporting daily contact with all students.

Supplementary Figure S3 shows the cumulative proportion of close contacts identified by questionnaire and UWB under different thresholds, counting backward from the final survey day. The total number of unique contacts identified by UWB at 1.5 m for ≥10 min over the full five-day period was set as the 100% reference. Questionnaire-based detection increased over the most recent two days but plateaued after the third day, suggesting limited recall of earlier interactions. In contrast, UWB-based detection—especially under the 1.5 m/10 min threshold—continued to accumulate contacts steadily across all five days. The official 1.0 m/15 min threshold consistently identified fewer contacts overall.

Supplementary Figure S4 illustrates consistency rates plotted against the number of identified close contacts under alternative thresholds. Panel A (UWB at 1.0 meter for ≥15 min, the official definition) shows a positive trend, indicating that participants who reported more contacts tended to have higher consistency rates—suggesting reduced discrepancy between questionnaire and UWB at higher contact counts. Panel B (UWB at 1.5 m for ≥10 min, the optimal threshold from sensitivity analysis) demonstrates that participants with larger numbers of close contacts identified by UWB were less accurately captured by questionnaires, suggesting that recall was particularly limited among individuals with many interactions. Panel C compares questionnaire-based reporting at 1.0 m for ≥15 min with UWB at 1.5 m for ≥10 min, showing that substantial overreporting was largely driven by a small number of individuals with very high reported contacts, such as the teacher, whereas for most participants, discrepancies were modest.

## Discussion

This study is the first to quantify discrepancies between self-reported and objectively measured close contacts in a real-world school environment using high-precision UWB positioning. Across five days of classroom observation, we found that the official Japanese definition of a close contact—within 1 m for at least 15 min—did not align well with actual interaction patterns, as measured by UWB. These discrepancies persisted despite the fixed-seating arrangement, which should have facilitated recall.

In our classroom setting, most interpersonal distances clustered around 2–3 m, while interactions within 1 meter—the basis for the official close contact definition—were relatively rare. This structural mismatch suggests that the current 1.0 meter for ≥15 min criterion may not adequately capture actual interaction patterns in schools. As a result, discrepancies between self-reported and UWB-measured contacts were frequent, with both underreporting (UWB > Questionnaire) and overreporting (Questionnaire > UWB) observed across participants. Revisiting operational definitions with reference to empirical distance distributions could reduce misclassification and improve the validity of contact tracing data.

Threshold calibration was a novel contribution of this work. Sensitivity analysis showed that the smallest discrepancies occurred at 1.5 m for ≥10 min. This threshold does not necessarily reflect a biologically optimal cut-off for transmission risk; rather, it appears to represent the point at which retrospective recall most closely aligns with objectively measured interactions. This distinction is critical: optimizing definitions for memory recall is methodologically useful, but such thresholds should not be interpreted as inherently epidemiologically meaningful.

The observed patterns are consistent with known recall biases. Overreporting may arise when participants overestimate the frequency or duration of brief interactions, especially under fixed-seating conditions where routine-based assumptions can blur recall. Conversely, underreporting may result from memory decay, particularly when the perceived consequences of contact designation are minimal—as in the post-pandemic context of this study. Reciprocity analysis revealed further asymmetry: many pairs disagreed on whether contact occurred, highlighting the subjectivity of memory-based assessments.

These findings have practical implications. Overreliance on retrospective self-reports can lead to substantial misclassification in contact tracing, with downstream operational consequences. Questionnaire-based identification increased during the two most recent days but plateaued after the third day, reinforcing concerns about memory degradation in multi-day recall. This pattern suggests that participants were more likely to recall recent interactions, whereas earlier contacts were more likely to be forgotten or unreported. In real-world settings, this may result in delayed or inaccurate contact identification, ultimately weakening containment efforts.

In our study, the teacher—who had frequent contact with all students—appeared to contribute disproportionately to overreporting, as suggested by stratified discrepancy patterns. However, given that only one teacher was included, this observation should be interpreted with caution and is not generalizable. Nonetheless, it highlights how high-contact individuals may inflate reported contact counts under memory-based systems, potentially leading to unnecessary quarantines and disruption in educational settings.

This concern is supported by prior modeling studies, which have shown that even modest delays in contact identification can significantly reduce the effectiveness of tracing efforts [22] and that the choice of testing strategy can dramatically influence the number of individuals placed under quarantine [23]. Large-scale surveillance data from Okinawa, Japan, further echo these concerns: classes were closed entirely upon the detection of a single case, despite an average of fewer than one secondary infection per event [24]. Collectively, these findings emphasize the importance of aligning contact tracing protocols not only with epidemiological risk, but also with practical accuracy and social feasibility.

Importantly, this study does not claim that UWB positioning is a gold standard for defining infectious exposures. Actual transmission risk depends on multiple factors—including ventilation, mask use, viral load, and host susceptibility—that were beyond the scope of this study. Rather, our goal was to assess whether current contact definitions align with real-world behavior and memory, and whether objective measurements can help refine these definitions for practical use.

This study has several limitations. First, measurements were confined to a single classroom in one junior high school, and lessons held outside the homeroom were excluded. These constraints limit the generalizability of our findings to more dynamic settings such as workplaces, public events, or other educational environments with variable seating and movement patterns. Second, although the UWB system provides nominal spatial accuracy of 10–50 cm, we did not empirically validate this accuracy under classroom-specific conditions. In particular, our analysis did not account for participant orientation or body-shadowing effects, which are known to affect signal quality. Deep learning–based models have shown promise in correcting for antenna orientation [25], and inertial measurement units (IMUs) can mitigate shadowing artifacts [26], but these enhancements were not implemented in our setup.

Third, the questionnaire data relied on retrospective recall across five consecutive days. This design inherently exposes responses to memory decay and routine-based assumptions, especially in a fixed-seating context. Indeed, the plateauing of new contacts after three days suggests diminishing recall reliability over time [27]. Fourth, we did not assess epidemiological outcomes such as secondary infections, and therefore cannot determine whether either method better predicts biologically meaningful exposure. Finally, unmeasured contextual factors—such as unrecorded interactions outside classroom hours—may have led to under-detection in both data sources.

Future studies should integrate UWB-based proximity data with microbiological or epidemiological outcomes to determine transmission-relevant contact thresholds. Such approaches may be particularly useful in healthcare settings, where contact-mediated transmission of pathogens—including multidrug-resistant organisms (MDROs)—poses significant challenges. For example, combining UWB-based spatiotemporal mapping with real-time diagnostic testing—such as PCR, LAMP, or antigen-based swab tests—could enable the direct validation of proximity-based definitions against confirmed transmission events. This would support the empirical calibration of close contact thresholds, moving beyond retrospective or observational assumptions. Current operational definitions of close contact, such as “within 1 meter for ≥15 minutes,” were largely derived from outbreak investigations during the SARS and MERS epidemics and adopted pragmatically by WHO and CDC in the early phase of the COVID-19 pandemic [28, 29, 3]. These thresholds were informed by observational data and expert consensus, such as droplet transmission studies and meta-analyses indicating increased infection risk at distances <1–2 meters, rather than through standardized experimental validation. These criteria often fail to account for contextual factors such as mask usage, ventilation, and directional orientation. By combining sensor-based data with pathogen detection, future research could contribute to the evidence-based redefinition of close contact thresholds and improve the accuracy of infection control policies in both educational and clinical environments.

In conclusion, this study provides real-world evidence that self-reported close contacts and UWB-measured contacts diverge substantially in a school environment. The discrepancies were driven not only by limitations in recall, but also by the structural mismatch between physical interactions and the operational definition of close contact. While 1.5 m for ≥10 min may offer a better balance between recall and measurement, it should not be mistaken for an epidemiologically validated threshold. Integrating objective spatiotemporal data with empirically calibrated definitions may support more accurate, efficient, and context-sensitive contact tracing while minimizing unnecessary social and educational disruption.

## Contributors

HS conducted the field survey, performed the data analyses, and was primarily responsible for generating the figures and tables. He also engaged in repeated discussions with KM regarding the interpretation of the data visualizations, contributing substantially to the refinement of key findings.

YO served as the second author and contributed critically to the methodological validation, particularly ensuring the robustness and reliability of the UWB positioning analyses described in the Methods section.

KM conceived the study, designed the questionnaire, secured the study sites, participated in the field survey, and drafted the initial manuscript. He also obtained and managed the research funding that supported this study, supervised the overall research process, and coordinated communication among collaborators and study sites.

The fourth through sixth authors are listed alphabetically, as they contributed equally and in comparable ways to the study. The final author order may be revised based on contributions during the revision process.

## Data sharing

The individual-level UWB positioning data and questionnaire responses contain potentially identifiable information and will be used in ongoing graduate student research projects, including network analyses. For this reason, the full dataset cannot be shared publicly at this time. Deidentified summary statistics and analytic code supporting the findings of this article will be made available from the corresponding author upon publication. The full deidentified dataset will be deposited in a public repository within 2 years of publication. All authors had full access to the data and accept responsibility for the decision to submit for publication.

## Declaration of interests

We declare no competing interests.

## Fund

KM acknowledges support from the Japan Society for the Promotion of Science (JSPS) KAKENHI [Grant 20H03940 and 20KK0367].The funding source had no role in study design, data collection, data analysis, interpretation, writing of the manuscript, or the decision to submit for publication. All authors had full access to the data in the study and accept responsibility for the decision to submit for publication.

## Acknowledgments

The authors appreciate the following institutions and individuals for their contributions to this study: Minabe Town Kamiminabe Junior High School, Principal Naomi Ogi, and Mr. Nakamura Kiichi, the homeroom teacher, whose cooperation was essential for conducting the study. In addition to supporting classroom-based data collection, they facilitated communication with participants and guardians, including distributing consent forms, providing explanations, and collecting signed agreements.

## Declaration of generative AI and AI-assisted technologies in the writing process

During the preparation of this work, generative AI tools, including OpenAI’s ChatGPT, were used to improve the clarity and readability of the manuscript under the full oversight of the authors. All content was reviewed and edited by the authors, who take full responsibility for the final content of the publication.

## Appendix

### Additional file 1

**Table S1. Summary of questionnaire respondents (N = 33)**

**Table S2. Parameters of the Weibull distribution fitted to pairwise distance data (all participants, five days combined)**

**Table S3. Daily Number of Close Contacts per Participant Identified by UWB and Questionnaire**

**Table S4. Daily Discrepancies Between UWB- and Questionnaire-Identified Close Contacts (1.0 m,** ≥**15 min Threshold)**

**Table S5. Discrepancies Under Alternative Distance–Time Thresholds for Close Contact Definition**

**Table S6. Daily Discrepancies Under the Optimal Threshold (1.5 m,** ≥**10 min)**

**Figure S1. Distribution of one-second pairwise distances between all participant pairs during the full survey period**

Distances were measured by UWB positioning at 1 Hz. The distribution shows a single peak at 2–3 m with a long right tail.

**Figure S2. Pairwise consistency rates based on questionnaires, by day**

Each box plot represents the median, IQR, and outliers for all pairs. Days are labeled Monday through Friday, corresponding to Day 1 through Day 5 in the main text. Teacher–student pairs showed zero agreement on all days.

**Figure S3. Cumulative proportion of close contacts identified by questionnaire and UWB, counting backward from the final survey day**

The figure shows the cumulative proportion of unique close contacts identified under three different methods: questionnaire (solid line), UWB at 1.0 m for ≥15 min (dashed line), and UWB at 1.5 m for ≥10 min (dotted line). The total number of contacts detected by UWB at 1.5 m/10 min over the five-day period is set as the 100% reference. The x-axis represents the number of days counting backward from Day 5, illustrating how recall and detection dynamics vary over time.

**Figure S4. Consistency rates versus actual contact counts under alternative thresholds.**

(A) UWB at 1.0 m for ≥15 min (official definition),

(B) UWB at 1.5 m for ≥10 min (optimal threshold from sensitivity analysis),

(C) Questionnaire (1.0 m, ≥15 min) vs. UWB (1.5 m, ≥10 min).

Panel A shows the relationship between consistency rates and the number of close contacts when UWB was evaluated at 1.0 m for ≥15 min (the official definition). A negative trend is observed, indicating that questionnaires tended to overreport contacts relative to UWB.

Panel B shows the same relationship when UWB was evaluated at 1.5 m for ≥10 min (the optimal threshold identified in sensitivity analyses). Here, participants with larger numbers of contacts were less consistently captured by questionnaires, suggesting that recall was particularly limited among highly interactive individuals.

Panel C compares questionnaire-based reporting at 1.0 m for ≥15 min with UWB at 1.5 m for ≥10 min. Overreporting was most pronounced among a small number of individuals with high self-reported contact counts. Notably, the teacher exhibited substantial negative discrepancies; however, given that only one teacher was included, this observation should be interpreted with caution and cannot be generalized. For most participants, discrepancies were relatively modest.

Positive values indicate more contacts detected by UWB (underreporting in questionnaires), whereas negative values indicate more contacts reported in questionnaires (overreporting).

Small red dots represent individual participants’ consistency rates for each day. The dashed line represents the fitted linear trend; the gray shaded area shows the 95% confidence interval.

### Additional file 2

**Questionnaire Survey (English Translation)**

## Notes

### Competing Interest Statement

The authors have declared no competing interest.

### Author Declarations

The study protocol was reviewed and approved by the Ethics Committee of Kyoto University Graduate School of Medicine (Approval No. R6-G-8-2).

## Reference

1. WHO. Contact tracing in the context of COVID-19. Interim guidance. Geneva: WHO; 2021.

2. Chu DK, Akl EA, Duda S, Solo K, Yaacoub S, Schünemann HJ; COVID-19 Systematic Urgent Review Group Effort (SURGE) study authors. Physical distancing, face masks, and eye protection to prevent person-to-person transmission of SARS-CoV-2 and COVID-19: a systematic review and meta-analysis. Lancet. 2020;395(10242):1973–87. doi:10.1016/S0140-6736(20)31142-9

3. National Institute of Infectious Diseases (Japan). Provisional guidelines for active epidemiological investigation for COVID-19 (Jan 17 and Jan 21, 2020 versions). Available from: https://id-info.jihs.go.jp

4. National Institute of Infectious Diseases (Japan). Provisional guidelines for active epidemiological investigation for COVID-19 (Apr 20, 2020 version). Available from: https://id-info.jihs.go.jp

5. National Institute of Infectious Diseases (Japan). Q&A on the revision of the definition of close contacts in the provisional guidelines for active epidemiological investigation (Apr 27, 2020). Available from: https://id-info.jihs.go.jp

6. Bernard HR, Killworth PD, Sailer L. Informant accuracy in social network data IV: an experimental attempt to predict actual communication from recall data comparison of clique-level structure in behavioral and cognitive network data. Soc Networks. 1982;11(1):30–66. doi: 10.1016/0049-089X(82)90006-0

7. Simons DJ, Chabris CF. What people believe about how memory works: a representative survey of the U.S. population. PLoS One. 2011;6(8):e22757. doi: 10.1371/journal.pone.0022757.

8. Voirin N, Payet C, Barrat A, Cattuto C, Khanafer N, Régis C, et al. Combining high-resolution contact data with virological data to investigate influenza transmission in a tertiary care hospital. Infect Control Hosp Epidemiol. 2015;36(3):254–60. doi: 10.1017/ice.2014.53.

9. Hornbeck T, Naylor D, Segre AM, Thomas G, Herman T, Polgreen PM. Using sensor networks to study the effect of peripatetic healthcare workers on the spread of hospital-associated infections. J Infect Dis. 2012;206(10):1549–57. doi: 10.1093/infdis/jis542.

10. Stopczynski A, Sekara V, Sapiezynski P, Cuttone A, Madsen MM, Larsen JE, Lehmann S. Measuring large-scale social networks with high resolution. PLoS One. 2014;9(4):e95978. doi: 10.1371/journal.pone.0095978.

11. Aiello AE, Simanek AM, Eisenberg MC, Walsh AR, Davis B, Volz E, et al. Design and methods of a social network isolation study for reducing respiratory infection transmission: The eX-FLU cluster randomized trial. Epidemics. 2016;15:38–55. doi: 10.1016/j.epidem.2016.01.001.

12. Salathé M, Kazandjieva M, Lee JW, Levis P, Feldman MW, Jones JH. A high-resolution human contact network for infectious disease transmission. Proc Natl Acad Sci U S A. 2010;107(51):22020–5. doi: 10.1073/pnas.1009094108.

13. Stehlé J, Voirin N, Barrat A, Cattuto C, Colizza V, Isella L, Régis C, Pinton JF, Khanafer N, Van den Broeck W, Vanhems P. Simulation of an SEIR infectious disease model on the dynamic contact network of conference attendees. BMC Med. 2011;9:87. doi: 10.1186/1741-7015-9-87.

14. Grantz KH, Lee EC, D’Agostino McGowan L, Lee KH, Metcalf CJE, Gurley ES, Lessler J. Maximizing and evaluating the impact of test-trace-isolate programs: A modeling study. PLoS Med. 2021;18(4):e1003585. doi: 10.1371/journal.pmed.1003585.

15. Qian L, Chan A, Cai J, Lewicke J, Gregson G, Lipsett M, et al. Evaluation of the accuracy of a UWB tracker for in-home positioning for older adults. Med Eng Phys. 2024;126:104155. doi: 10.1016/j.medengphy.2024.104155.

16. Alarifi A, Al-Salman A, Alsaleh M, Alnafessah A, Al-Hadhrami S, Al-Ammar MA, et al. Ultra wideband indoor positioning technologies: analysis and recent advances. Sensors (Basel). 2016;16(5):707. doi:10.3390/s16050707

17. Gezici S, Poor HV, Sahinoglu Z, Kobayashi H, Molisch AF, et al. Localization via ultra-wideband radios: a look at positioning aspects for future sensor networks. IEEE Signal Process Mag. 2005;22(4):70–84. doi:10.1109/MSP.2005.1458289

18. Shinto H, Ohki Y, Mizumoto K, Saito K. Visualization of interpersonal communication using indoor positioning technology with UWB tags. Manuscript under peer review.

19. Xu YS, Shimakawa Y, Chowell G, Matsuyama R, Nagamoto T, Omori R, et al. Determinants of COVID-19 Outbreak Size in Elderly Residential Facilities in Okinawa Prefecture, Japan, April to June 2022. Manuscript under peer review.

20. Ministry of Education, Culture, Sports, Science and Technology (MEXT). School education system in Japan. Tokyo: MEXT; 2020.

21. Ministry of Education, Culture, Sports, Science and Technology (MEXT). Regarding the implementation of the five-day school week system. Tokyo: MEXT; 2002. Available from: https://www.mext.go.jp

22. Kretzschmar ME, Rozhnova G, van Boven M. Impact of delays on effectiveness of contact tracing strategies for COVID-19: a modelling study. Lancet Public Health. 2020;5(8):e452–9. doi:10.1016/S2468-2667(20)30157-2

23. Quilty BJ, Clifford S, Hellewell J, Russell TW, Kucharski AJ, Flasche S, et al. Quarantine and testing strategies in contact tracing for SARS-CoV-2: a modelling study. Lancet Public Health. 2021;6(3):e175–83. doi:10.1016/S2468-2667(20)30308-X

24. Takayama Y, Shimakawa Y, Matsuyama R, Chowell G, Omori R, Nagamoto T, Yamamoto T, Mizumoto K. SARS-CoV-2 infection in school settings, Okinawa Prefecture, Japan, 2021–2022. Emerg Infect Dis. 2024;30(11):2343–51. doi:10.3201/eid3011.240638

25. Urwan S, Cwalina KK. User orientation detection in relation to antenna geometry in ultra-wideband wireless body area networks using deep learning. Sensors (Basel). 2024;24(7):2060. doi:10.3390/s24072060

26. De Cock C, Tanghe E, Joseph W, Plets D. Robust IMU-based mitigation of human body shadowing in UWB indoor positioning. Sensors (Basel). 2023;23(19):8289. doi:10.3390/s23198289

27. Bernard HR, Killworth PD, Kronenfeld D, Sailer L. The problem of informant accuracy: the validity of retrospective data. Annu Rev Anthropol. 1984;13:495–517. doi:10.1146/annurev.an.13.100184.002431

28. World Health Organization. Global Surveillance for COVID-19: Interim Guidance. 2020. Available from: https://www.who.int/docs/default-source/coronaviruse/2020-03-20-surveillance.pdf [accessed Sept 1, 2025].

29. Centers for Disease Control and Prevention. Case Surveillance Definition for COVID□19. 2021.Available from: https://ndc.services.cdc.gov/case-definitions/coronavirus-disease-2019-2021/ [accessed Sept 1, 2025].

