## Supplementary file 1 for "Discrepancies Between Self-Reported and High-Precision UWB-Measured Close Contacts: Evidence from a Japanese Junior High School"

### Additional file 1:

1. Supplementary Methods
2. Supplementary Tables S1-S2

Table S1. Summary of questionnaire respondents (N = 33)

Table S2. Parameters of the Weibull distribution fitted to pairwise distance data (all participants, five days combined)

Table S3. Daily Number of Close Contacts per Participant Identified by UWB and Questionnaire.

Table S4. Daily Discrepancies Between UWB- and Questionnaire-Identified Close Contacts (1.0 m, ≥15 min Threshold) .

Table S5. Discrepancies Under Alternative Distance–Time Thresholds for Close Contact Definition.

Table S6. Daily Discrepancies Under the Optimal Threshold (1.5 m, ≥10 min).

3. Supplementary Figure S1-S4

Figure S1. Distribution of one-second pairwise distances between all participant pairs during the full survey period.

Figure S2. Pairwise consistency rates based on questionnaires, by day.

Figure S3. Cumulative proportion of close contacts identified by questionnaires and UWB, counting backward from the final survey day.

Figure S4. Consistency rates versus actual contact counts under alternative thresholds.

**Supplementary Methods**

**Consistency rate**

The consistency rate was calculated by comparing the presence or absence of close contacts for each participant on each survey day, based on data from both the questionnaire and UWB methods. For each day, two 33×33 binary matrices were constructed to represent pairwise close contacts, one from questionnaire responses and the other from UWB measurements. Questionnaire data were symmetrized: if either member of a pair reported a close contact, both corresponding cells were marked as 1. UWB data were inherently bidirectional. The consistency matrix was generated by subtracting the questionnaire matrix from the UWB matrix. This yielded values of +1 (contact detected only by UWB = underreporting), –1 (only by questionnaire = overreporting), or 0 (detected by both methods). For each participant, the row-wise sum of the 33×33 matrix was computed, then divided by 32 (i.e., the number of possible counterparts), yielding a daily consistency rate per participant. Positive consistency rates indicate that UWB identified more contacts than questionnaires (suggesting underreporting), while negative values indicate more contacts reported in questionnaires (suggesting overreporting). This process was repeated for each of the five survey days, resulting in 165 participant-day observations.

| **Table S1. Summary of questionnaire respondents (N = 33)** | | | |
| --- | --- | --- | --- |
|  |  | Mean (SD) or n (%) | |
| Role | Student | 32 | (97.0) |
|  | Teacher | 1 | (3.0) |
| Gender (student) | Male | 18 | (56.2) |
|  | Female | 14 | (43.8) |
| Gender (teacher) | Male | 1 | (100) |
| Confidence level | 1 (Lowest) | 1 | (3.1) |
|  | 2 | 6 | (18.8) |
|  | 3 | 7 | (21.9) |
|  | 4 | 14 | (43.8) |
|  | 5 (Highest) | 4 | (12.5) |

Two participants who carried UWB tags did not return the questionnaire. Values are shown as mean (SD) or n (%). Categories include gender, role (student or teacher), and self-reported confidence in recall (5-point Likert scale). Percentages may not sum to 100 due to rounding. Total number of respondents: 33.

Table S2. Parameters of the Weibull distribution fitted to pairwise distance data (all participants, five days combined)

| Parameter | Estimate | 95% CI |
| --- | --- | --- |
| Shape (k) | 2.154 | 2.153 - 2.155 |
| Scale (λ) | 3.601 | 3.604 - 3.606 |

Parameters were estimated by maximum likelihood in Python. The Weibull distribution provided the best fit compared with alternative distributions (exponential, log-normal) based on Akaike information criterion (AIC).

Table S3. Daily Number of Close Contacts per Participant Identified by UWB and Questionnaire

| Number of close contacts | | | | | | |  |
| --- | --- | --- | --- | --- | --- | --- | --- |
| UWB | | | | | | |  |
|  | Monday  (Day 1) | Tuesday  (Day 2) | Wednesday  (Day 3) | Thursday  (Day 4) | Friday  (Day 5) | All daily observations combined | Five-Day Cumulative Contacts per Participant (Unique Individuals) |
| Mean | 3.3 | 2.4 | 0.5 | 2.4 | 2.8 | 2.3 | 5.7 |
| Median | 4.0 | 3.0 | 0.0 | 2.0 | 3.0 | 2.0 | 6.0 |
| IQR | 0.0, 5.0 | 0.0, 4.0 | 0.0, 1.0 | 0.0, 4.0 | 0.25, 4.8 | 0.0, 4.0 | 4.0, 7.0 |
| Questionnaire | | | | | | |  |
|  | Monday  (Day 1) | Tuesday  (Day 2) | Wednesday  (Day 3) | Thursday  (Day 4) | Friday  (Day 5) | All daily observations combined | Five-Day Cumulative Contacts per Participant (Unique Individuals) |
| Mean | 7.7 | 8.4 | 9.6 | 9.5 | 9.3 | 8.9 | 13.5 |
| Median | 8.0 | 8.5 | 9.0 | 9.0 | 9.0 | 9.0 | 14.0 |
| IQR | 0.5, 11.0 | 2.3, 12.8 | 5.0, 14.8 | 4.0, 14.8 | 4.0, 14.8 | 4.0, 13.7 | 9.0, 17.0 |

Summary statistics (mean, median, and IQR) of daily close contact counts over five survey days, identified separately by UWB positioning and self-reported questionnaires.

All daily observations combined show all close contacts (maximum 165 data points) of each participant for each day over the week. Summary shows the total number of close contacts for each participant over the week.

Data corresponds to Figure 1.

Table S4. Daily Discrepancies Between UWB- and Questionnaire-Identified Close Contacts (1.0 m, ≥15 min Threshold)

| Discrepancy ( UWB- questionnaire, consistency rate) | | | | | | |
| --- | --- | --- | --- | --- | --- | --- |
|  | Monday  (Day 1) | Tuesday  (Day 2) | Wednesday  (Day 3) | Thursday  (Day 4) | Friday  (Day 5) | All daily observations combined |
| Mean | −0.12 | −0.17 | −0.26 | −0.19 | −0.19 | −0.19 |
| Median | −0.11 | −0.12 | −0.24 | −0.16 | −0.16 | −0.16 |
| IQR | −0.19, 0.00 | − 0.25, −0.03 | − 0.36, − 0.13 | −0.30, −0.08 | −0.27, − 0.07 | −0.28, −0.05 |

Daily consistency rate statistics (mean, median, IQR) comparing UWB and questionnaire-based contact identification under the official 1.0 m, ≥15 min definition. Corresponds to Figure 2.

Table S5. Discrepancies Under Alternative Distance–Time Thresholds for Close Contact Definition

| Discrepancy ( UWB- questionnaire, consistency rate) | | | | | | |
| --- | --- | --- | --- | --- | --- | --- |
| Threshold | | | | | | |
|  | 1.0m, 15min | 1.0m, 10min | 1.0m, 5min | 1.5m, 15min | 1.5m, 10min | 1.5m, 5min |
| Mean | −0.20 | −0.17 | −0.09 | −0.11 | −0.05 | 0.07 |
| Median | −0.17 | −0.11 | −0.03 | −0.06 | −0.00 | 0.09 |
| IQR | −0.29, −0.06 | −0.26, −0.03 | −0.17, 0.00 | −0.20, 0.00 | −0.14, 0.09 | 0.00, 0.23 |

Comparison of consistency rate statistics across six distance–time thresholds (1.0 m / 1.5 m × 5, 10, 15 min). Used to identify the threshold minimizing discrepancies. Corresponds to Figure 3.

Table S6. Daily Discrepancies Under the Optimal Threshold (1.5 m, ≥10 min)

| Discrepancy ( UWB- questionnaire, consistency rate) | | | | | | |
| --- | --- | --- | --- | --- | --- | --- |
|  | Monday  (Day 1) | Tuesday  (Day 2) | Wednesday  (Day 3) | Thursday  (Day 4) | Friday  (Day 5) | All daily observations combined |
| Mean | 0.03 | −0.02 | −0.19 | −0.07 | 0.00 | −0.05 |
| Median | 0.05 | 0.00 | −0.16 | 0.00 | 0.01 | 0.00 |
| IQR | 0.00, 0.11 | −0.06, 0.08 | −0.28, −0.08 | −0.14, 0.03 | −0.03, 0.11 | −0.14, 0.08 |

Daily summary statistics of consistency rates using the optimal threshold (1.5 m for ≥10 minutes), which minimized discrepancies between UWB and questionnaire methods. Corresponds to Figure 4.

Figure S1. Distribution of one-second pairwise distances between all participant pairs during the full survey period

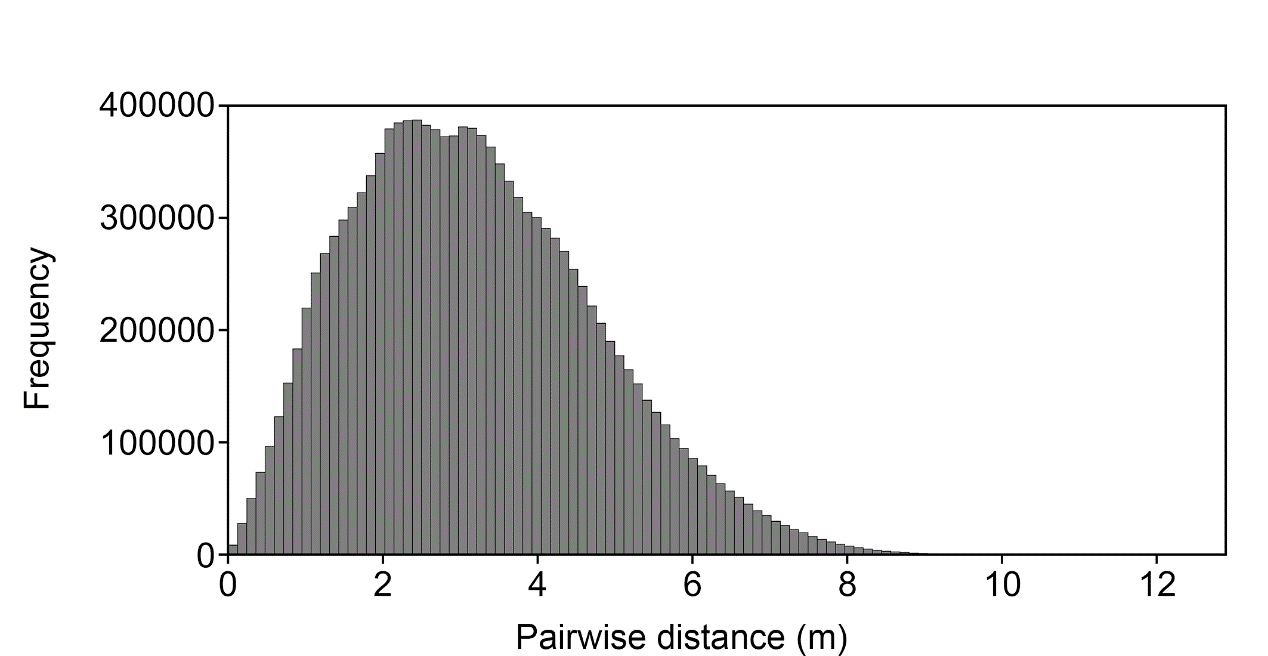

Figure S2. Pairwise consistency rates based on questionnaires, by day

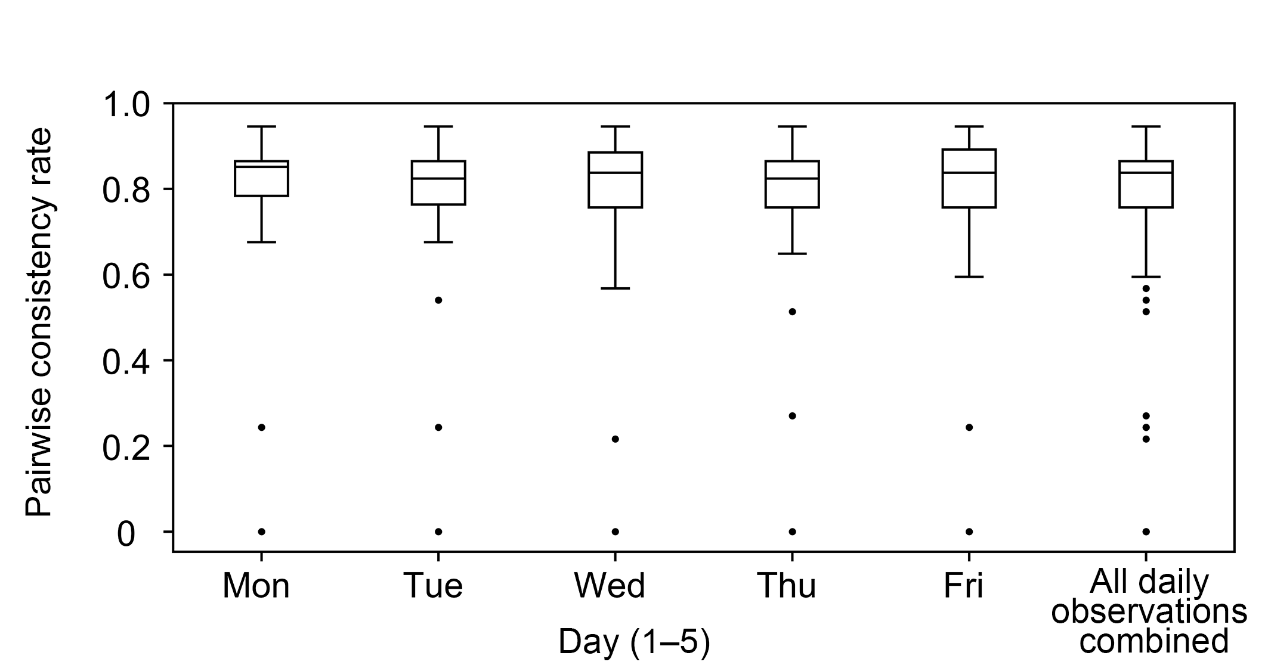

Figure S3. Cumulative proportion of close contacts identified by questionnaires and UWB, counting backward from the final survey day.

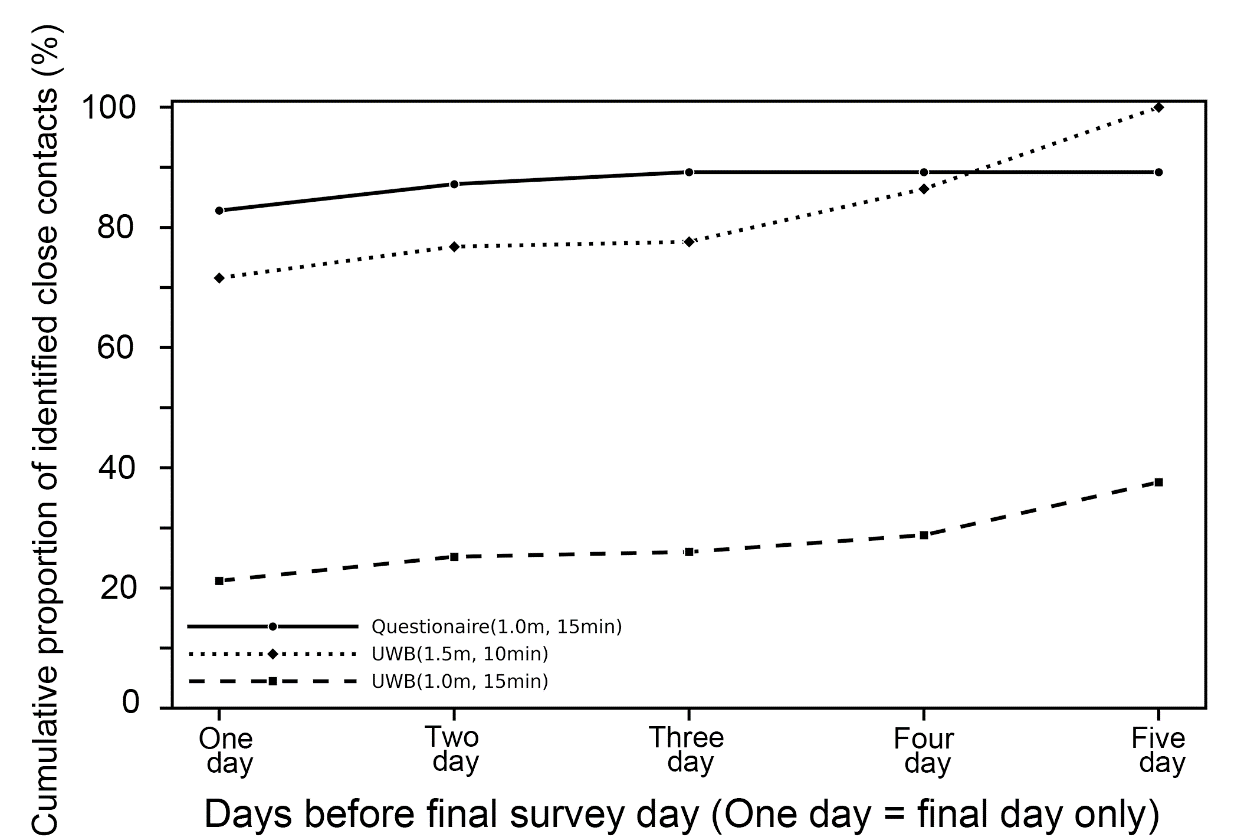

Figure S4. Consistency rates versus actual contact counts under alternative thresholds.

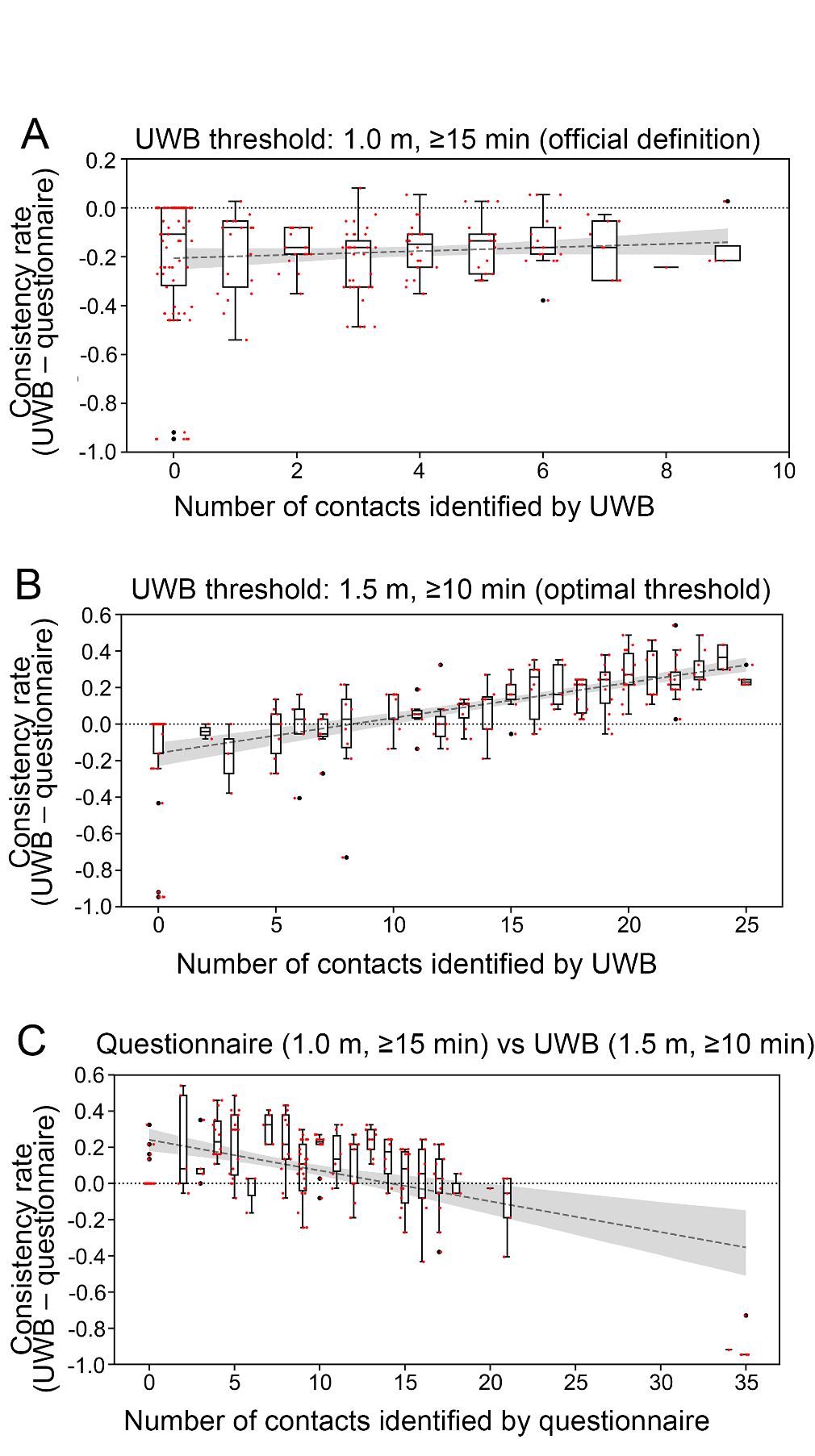
