## Supplementary file 2 for "Discrepancies Between Self-Reported and High-Precision UWB-Measured Close Contacts: Evidence from a Japanese Junior High School"

For School, For teachers/students

### Questionnaire Survey (English Translation)

**Date: (Year)  2025  (Month)    (Day)**

**Q1. Please enter your attendance number (or your tag number if you are a teacher).**

**(          )**

**Q2. Please indicate your grade and class (or your assigned class if you are a teacher).**

**(Grade, Class)**

**Q3. Please indicate your sex (check one)**

**Male・Female・Others**

**Q4. Please indicate your age.**

**(   ) years old**

**Q5. Please check your role.**

**Teacher / Student**

**Q6. For each day, please indicate individuals whom you consider having been "close contacts" during the target period and within the target area.**
**Definitions are provided below:**

- **"Close contact": Any individual who was within 1 meter for a cumulative total of at least 15 minutes, regardless of mask use.**
- **"Target period": The period during which you carried the tag each day (from homeroom or distribution to end-of-day return; Jan 20–24, 2025).**
- **"Target area" :**

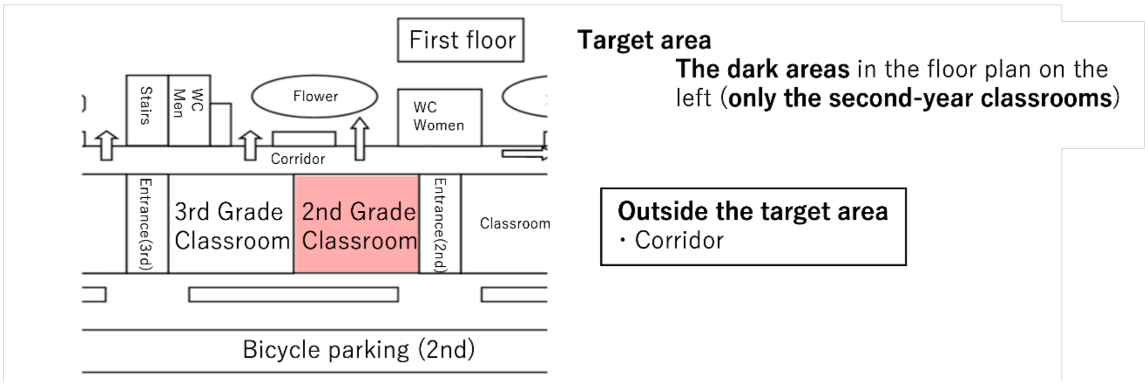

| Role | Grade | Number | 20 Jan (Mon) | 21 Jan (Tue) | 22 Jan (Wen) | 23 Jan (Thu) | 24 Jan (Fri) |
|---|---|---|---|---|---|---|---|
| Student | 2 | 1 | | | | | |
| | | 2 | | | | | |
| | | 3 | | | | | |
| | | 4 | | | | | |
| | | 5 | | | | | |
| | | 6 | | | | | |
| | | 7 | | | | | |
| | | 8 | | | | | |
| | | 9 | | | | | |
| | | 10 | | | | | |
| | | 11 | | | | | |
| | | 12 | | | | | |
| | | 13 | | | | | |
| | | 14 | | | | | |
| | | 15 | | | | | |
| | | 16 | | | | | |
| | | 17 | | | | | |
| | | 18 | | | | | |
| | | 19 | | | | | |
| | | 20 | | | | | |
| | | 21 | | | | | |
| | | 22 | | | | | |
| | | 23 | | | | | |
| | | 24 | | | | | |
| | | 25 | | | | | |
| | | 26 | | | | | |
| | | 27 | | | | | |
| | | 28 | | | | | |
| | | 29 | | | | | |
| | | 30 | | | | | |
| | | 31 | | | | | |
| | | 32 | | | | | |
| | | 33 | | | | | |
| | | 34 | | | | | |
| | | 35 | | | | | |
| | | 36 | | | | | |
| | | 37 | | | | | |
| Teacher | | 1 | | | | | |

**Q7. The list of close contacts you selected in Q6 will be compared with the list generated by the device. How confident are you that your memory-based selections match the device-based results? Please circle one:**

1. **Not confident at all**
2. **Slightly confident**
3. **Neutral**
4. **Confident**
5. **Very Confident**

**Q8. Free comment (Please let us know if you have any feedback or suggestions for improving this survey.)**

**This concludes with the questionnaire.**

**Before submitting, please check for any missing responses.**
**Thank you very much for your cooperation.**
